# Impact of January 2021 curfew measures on SARS-CoV-2 B.1.1.7 circulation in France

**DOI:** 10.1101/2021.02.14.21251708

**Authors:** Laura Di Domenico, Chiara E. Sabbatini, Giulia Pullano, Daniel Lévy-Bruhl, Vittoria Colizza

**Affiliations:** INSERM, Sorbonne Université, Pierre Louis Institute of Epidemiology and Public Health, Paris, France; Orange Labs, Sociology and Economics of Networks and Services (SENSE), Chatillon, France; Santé publique France, Saint-Maurice, France; Tokyo Tech World Research Hub Initiative, Institute of Innovative Research, Tokyo Institute of Technology, Tokyo, Japan

## Abstract

Facing B.1.1.7 variant, social distancing was strengthened in France in January 2021. Using a 2-strain mathematical model calibrated on genomic surveillance, we estimated that curfew measures allowed hospitalizations to plateau, by decreasing transmission of the historical strain while B.1.1.7 continued to grow. School holidays appear to have further slowed down progression in February. Without progressively strengthened social distancing, a rapid surge of hospitalizations is expected, despite the foreseen increase in vaccination rhythm.

---

The new SARS-CoV-2 B.1.1.7 variant (20I/501Y.V1, also called variant of concern VOC 202012/01) initially detected in the UK^1,2^ has rapidly expanded its geographical range to European countries^3^. A large-scale genome sequencing initiative conducted in France on January 7-8 (Flash1 survey^4,5^) reported that 3.3% of all SARS-CoV-2 detections were B.1.1.7 viruses. To limit SARS-CoV-2 spread, strengthened social distancing measures were implemented in the country in the month of January. Starting from a curfew at 8pm in mid-December, curfew was anticipated at 6pm in several departments since January 2, due to deteriorating indicators, then extended nationwide on January 16, with renewed recommendations on telework and preventive measures. On January 31, heightened controls of the respect of the measures and closure of large commercial centers were applied.

The presence of B.1.1.7 variant on the territory, however, poses critical challenges to epidemic control. Its higher transmissibility represents a strong selective advantage to rapidly become the dominant strain^1,2,6–9^. Social distancing has a differential impact on the variant and the historical strain, not visible in surveillance data monitoring variant frequency at specific-date surveys. Assessing the impact of implemented measures on the two strains through modeling is key for epidemic management.

## Modeling SARS-CoV-2 2-strain transmission dynamics

We extended a previously developed age-stratified transmission model that was used to assess the impact of interventions against COVID-19 pandemic in 2020 in France^10–12^, fitted to hospital admission data and validated against serological studies’ estimates^10^. The model is discrete, stochastic, and integrates demography, age profile, social contacts, mobility data over time to account for social distancing measures. Details are provided in Ref.^10^ and the Supplement.

The model was extended to describe the circulation of two SARS-CoV-2 variants – the historical strain and B.1.1.7. Variant circulation was initialized on Flash1 data^4^: France (3.3%); Île-de-France region, reporting the highest penetration (6.9%); Nouvelle Aquitaine region, reporting one of the lowest penetration (1.7%). Complete cross-immunity and 60% transmissibility increase (range: 50-70%) were considered based on available knowledge^1,2,5^.

The model was fitted to daily hospital admission data in each territory to evaluate the impact of curfew in January (w02-w05), and of curfew and school holidays in February (w06-w09, with regional calendars; w06-w07 in Nouvelle Aquitaine, w07-w08 in Île-de-France). We projected future trends in hospitalizations at the end of holidays assuming the estimated curfew conditions, along with the strengthening and relaxation of measures (corresponding to 15% reduction or increase of the effective reproductive number estimated for the curfew, respectively).

Vaccination prioritized to older age classes was simulated according to current daily rhythm of 100,000 doses/day^13^, increased to 200,000 (first) doses/day (accelerated rhythm) from w10 following recent government announcements^14^ (Supplement). An optimistic rhythm of 300,000 (first) doses/day from w10 was considered for sensitivity.

### Estimated impact of social distancing measures and resulting B.1.1.7 trends

After an increase registered in hospital admission data from December (average 6,700 weekly hospitalizations at national level) to early January (about 9,000 in w02), the epidemic plateaued in the second half of the month, following the progressive implementation of curfew. Based on the estimated B.1.1.7 prevalence on January 7-8 and the reported hospitalizations in w02-05, the model explains this plateau as the counterbalance between two opposing dynamics: a decreasing circulation of the historical strain (with effective reproductive numbers 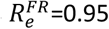 (95% confidence interval 0.94-0.96), 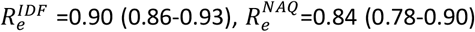 in w04 for France (FR), Île-de-France (IDF), Nouvelle Aquitaine (NAQ), respectively) opposed to the exponential increase of the variant. Curfew and other social distancing measures reduced the reproductive number of the historical strain below 1, but it was not enough to prevent the increasing B.1.1.7 dynamics (estimated *R*_*e*_ largely above 1 in all regions, 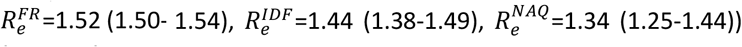. School holidays further slowed down the historical strain (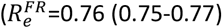 and 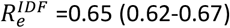 in w08, 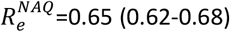 in w07), but were still insufficient against the variant (median *R*_*e*_ > 1 in all territories).

The projected increase of B.1.1.7 prevalence over time was confirmed by preliminary Flash2 survey^15^ data and more recent suspect B.1.1.7 data (i.e. not confirmed by sequencing, see Supplement; **Figure 2**). Data also matched estimated dominance dates, showing that B.1.1.7 accounted for the majority of cases by week 8 in France and Nouvelle Aquitaine, week 7 in Île-de-France. The variant is expected to increase by more than 55% the overall effective reproductive number by March 15 in Île-de-France, March 24 in France, April 3 in Nouvelle Aquitaine, compared to a situation without the variant.

**Figure 1.**
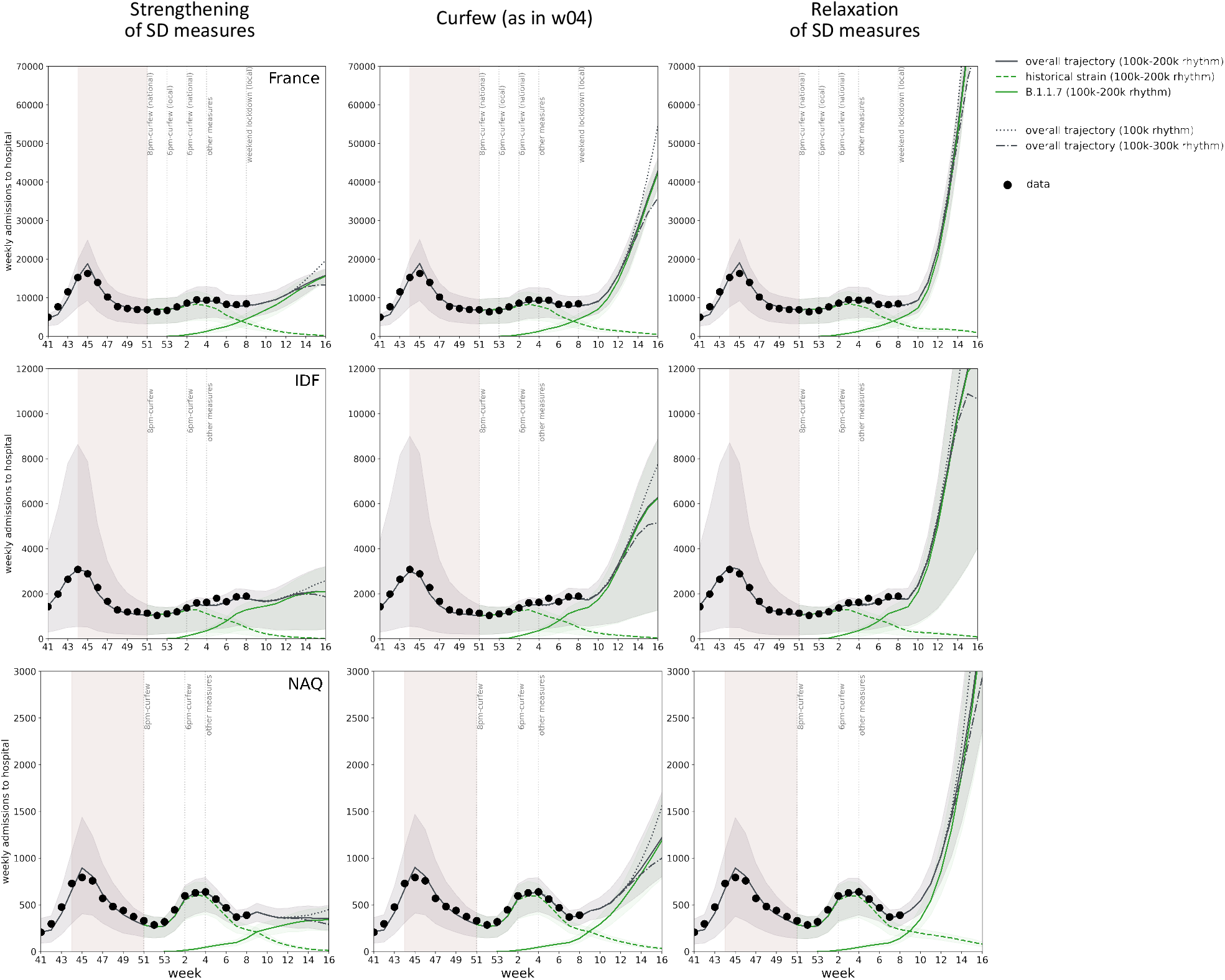
Projected weekly hospital admissions due to SARS-CoV-2 historical strain and B.1.1.7 variant in France and in the two regions under study. From left to right, different scenarios considered after winter school holidays: strengthening of social distancing (SD) measures scenario, equivalent to the second lockdown; curfew scenario, estimated in w04 and assuming no additional changes; relaxation of SD measures scenario, compatible with the situation at the start of the year before increased restrictions. From top to bottom: France, Île-de-France (IDF), Nouvelle Aquitaine (NAQ). The solid grey curve refers to the median overall trajectory, obtained under the accelerated vaccination rollout (100k-200k doses/day) and due to the concurrent circulation of the historical strain (dashed green curve) and B.1.1.7 variant (solid green curve), assuming 60% increase in transmissibility (50% and 70% increases are reported in the Supplement). A slower (100k, dotted curve) and optimistic (100k-300k, dot-dashed curve) vaccination rhythms are also shown (only median curves of the overall trajectories are shown, for the sake of visualization). The shaded area around the curves corresponds to the 95% probability range obtained from 500 stochastic simulations. Dots correspond to weekly hospital admission data. The model is fit to daily hospital admissions since the start of the epidemic, propagating uncertainty over time; the figure shows weekly data to simplify the visualization. The second wave is shown for reference, together with indications of the timing of social distancing measures; the shaded rectangle around the second wave corresponds to the second lockdown.

**Figure 2.**
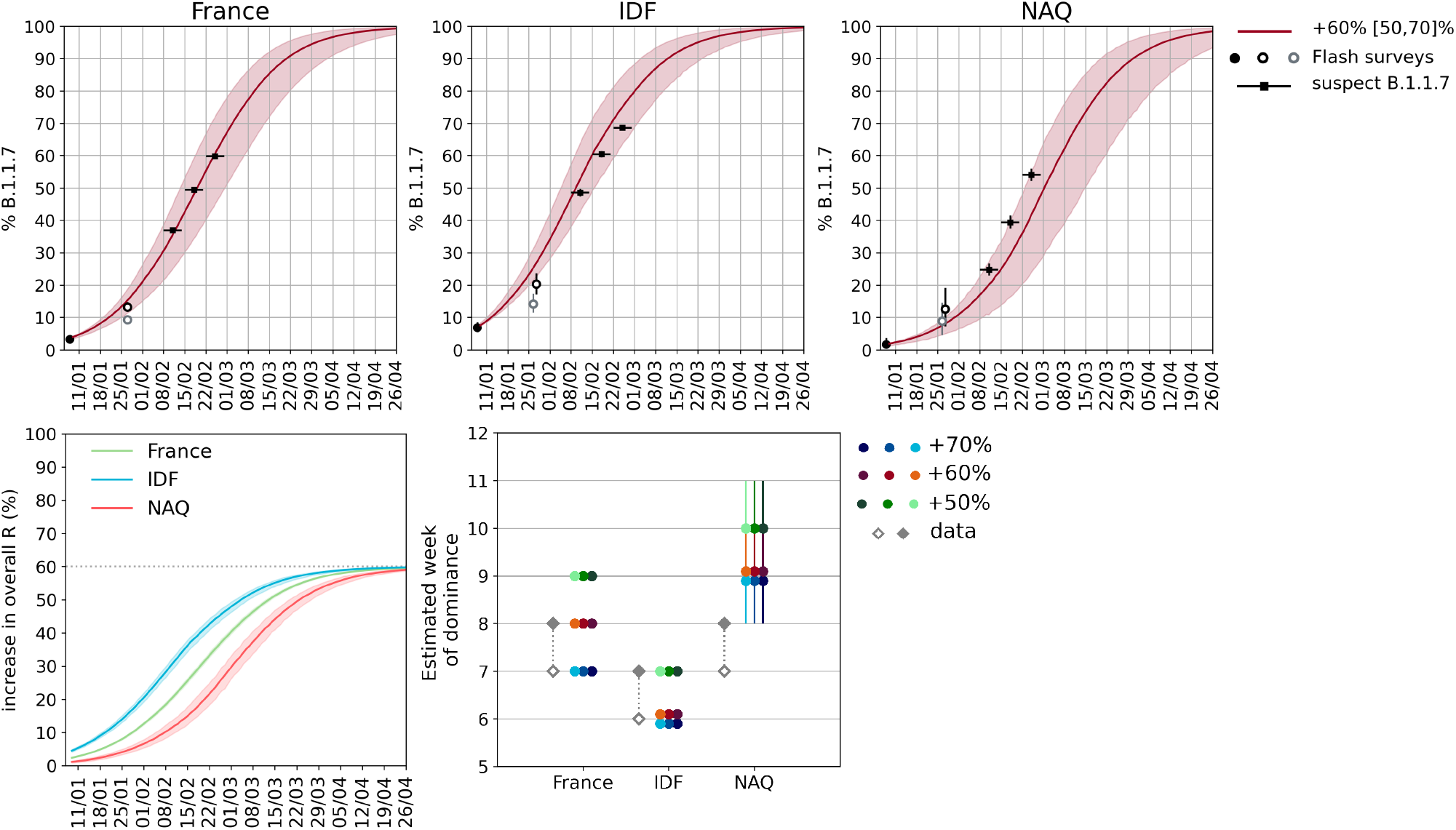
B.1.1.7 projected prevalence over time and estimated dominance week in France and in the two regions under study. Top: estimated percentage of B.1.1.7 cases over time, assuming 60% (50-70%) increase in transmissibility for the variant. From left to right: France, Île-de-France (IDF), Nouvelle Aquitaine (NAQ). Circles represent the estimates from Flash surveys. As sequencing is still ongoing, estimates from the second Flash survey are based on S-gene target failure percentages of positive tests for each territory and positive predicted value between 70% (grey border circle, national estimate from Flash1) and 100% (black border circles). Weekly data on suspect B.1.1.7 are reported with black squares; the horizontal line correspond to the week of reference. 95% confidence interval are estimated assuming a beta-binomial distribution to allow for dispersion due to spatial heterogeneity and variations in detection protocols. Bottom left: percentage increase in the overall effective reproductive number at the population level due to the increased penetration of the variant, assuming 60% increase in transmissibility of the variant. Curves represent median values; shaded areas around the curves represent 95% probability ranges obtained from 500 stochastic simulations. Bottom right: estimated week of B.1.1.7 dominance, assuming 60% (50-70%) increase in transmissibility for the variant, and considering the curfew scenario (middle point) and the scenarios with strengthening (lighter color, left point) and relaxation (darker color, right) of social distancing measures. Error bars represent 95% probability ranges. Grey diamonds correspond to the last week the reported frequency was <50% (void symbol), and the first week with reported frequency >50% (filled symbol).

### Projected hospitalizations under different scenarios

If the epidemic progresses under curfew conditions estimated before school holidays, and vaccination is accelerated, November peak hospitalization levels (close to hospital capacity in a number of regions) would be reached around w13 in France, w12 in Île-de-France, and w14 in Nouvelle Aquitaine (**Table 1**). The partial relaxation of social distancing – approximately corresponding to the epidemic conditions at the turn of the year before stricter measures were implemented – would anticipate these estimates of at least 1 week. Stronger social distancing, equivalent to the efficacy measured during the second lockdown, would maintain hospitalizations below the peak of the second wave in Île-de-France and Nouvelle Aquitaine. However, it may not be enough to avoid a third wave in France, even under the accelerated vaccination rhythm (100k-200k doses/day). Accelerated and optimistic vaccination rollouts would reduce weekly hospitalizations by about 20% and 35% in w16 compared to a stable vaccination campaign without acceleration.

**Table 1.** Estimated week at which hospitalizations exceed the peak value of the second wave in France and in the two regions under study. Projections after school holidays consider the curfew scenario (estimated before school holidays, assuming no additional interventions, central column), a scenario assuming a strengthening of social distancing (SD) measures (left column), and one assuming a relaxation of social distancing measures (right column). Results correspond to 50%, 60%, and 70% increase of transmissibility of the variant (60% being the reference value), and are obtained under the 100k-200k daily doses rhythm (see Supplement for details and results from other daily rhythms). Uncertainties refer to 95% probability ranges; “--” indicates that the peak level is not reached. Results cannot integrate yet the effect of the weekend lockdowns in certain areas, and do not include Easter school holidays.

|  |  | Peak weekly hospitalizations of second wave |  |  |  |
| --- | --- | --- | --- | --- | --- |
|  |  | B.1.1.7 transmissibility advantage | Strengthening of SD measures | Curfew (as in w04) | Relaxation of SD measures |
| France | 17,000 weekly hospitalizations | 50% increase | -- | week 14 (13-15) | week 13 (12-13) |
|  |  | 60% increase | week 16 (16-17) | week 13 (12-14) | week 12 (11-13) |
|  |  | 70% increase | week 14 (13-15) | week 12 (12-13) | week 12 (11-12) |
| Île-de-France | 3,000 weekly hospitalizations | 50% increase | -- | week 12 (12-13) | week 11 (10-13) |
|  |  | 60% increase | -- | week 12 (11-14) | week 11 (10-13) |
|  |  | 70% increase | -- | week 11 (11-14) | week 11 (10-13) |
| Nouvelle Aquitaine | 800 weekly hospitalizations | 50% increase | -- | after week 16 | week 13 (12-14) |
|  |  | 60% increase | -- | week 14 (12-16) | week 12 (11-13) |
|  |  | 70% increase | after week 16 | week 12 (11-14) | week 11 (11-12) |

## Discussion

We estimated that social distancing progressively implemented at the start of January 2021 was able to bring the effective reproductive number of the historical strain below 1, leading to its decline while B.1.1.7 cases exponentially increased. School holidays further slowed down the dynamics. The predicted growth in variant frequency and dominance date matched recent data.

Social distancing was the combined effect of imposed restrictions^16^ and individual response to renewed recommendations on telework and risk reduction. Telework, estimated from mobility data^10,17^, was maintained in January at the levels reached before releasing the second lockdown. Measures, however, were not enough to lead to a decline in the variant spread, not even under the additional impact of holidays, due to variant’s more efficient transmission.

Strengthening social distancing through a mild lockdown, as the one implemented to curb the second wave, would allow certain regions to avoid a third wave of the same magnitude of the second, due to acquired immunity (Île-de-France) or current conditions (Nouvelle Aquitaine, achieving a marked decrease in hospitalizations in January-February). Its duration would be longer than one month. Accelerating vaccination rollout is key^18^, but even optimistic rollout plans would lead to a rapid resurgence of cases under curfew only. Additional restrictive measures as the weekend lockdown recently implemented in high alert areas (Nice, Dunkerque) will contribute to decelerate such resurgence. Their effectiveness is expected to lie between the ones estimated for curfew and November mild lockdown. The latter, however, will unlikely prevent second peak-like hospitalization levels in April in France, even under the accelerated vaccination rollout. More rigorous and intensified social distancing over time should be anticipated to curb B.1.1.7 increasing pressure on viral circulation.

Our study has limitations. We could not yet include the impact of localized weekend lockdowns, and did not consider Easter school holidays, nor seasonal effects. All these elements are expected to contribute to slow down viral circulation. Results are based on estimated curfew impact and scenarios anticipating a possible strengthening or relaxation of social distancing. Changes of behaviors, as a progressive abandon of telework due to fatigue, or increased risk prevention due to growing concern, are not considered. Our analysis based on the estimated transmissibility advantage at the national level^5^ identifies differences between the two regions. These could be partly due to biases affecting Flash survey data and linked to reinforced tracing around suspected or confirmed variants, expected to be stronger in regions experiencing small-size epidemics (e.g. Nouvelle Aquitaine). We did not consider in the main analysis additional differences between the variant and the historical strain beyond the transmissibility advantage. The recently estimated increased hospitalization rate associated to B.1.1.7 infection^19^ would lead to a higher peak in the projected hospitalizations at the end of April (Supplement). Other variants were not considered here, as estimated at a lower penetration, but their circulation will likely concur to the expected case surge^20^.

## Data Availability

All data are available at the references cited.

## SUPPORTING INFORMATION

## 1. Surveillance data

### 1.1. Hospital surveillance data

Our model was fitted to daily hospital admission data to capture the epidemic trajectory over time (see the Inference section). This represents an exhaustive dataset of individuals affected with COVID-19 and requiring hospitalization. They are among the most robust data sources to use in COVID-19 epidemiological studies, as they are not affected by changes in detection and sampling, as it happens for detected cases, and do not suffer the strong delays or uncertainty in classification of the number of deaths. These data have been used throughout 2020 in France to respond to the health crisis^1–3^ and are also routinely used by local, national, and international health agencies for their assessments of the epidemiological situations. Data include admissions to conventional hospitalizations for COVID-19 by date of admission, and exclude transfer of patients and rehabilitation.

### 1.2. Genomic surveillance data

Two large-scale genome sequencing initiatives (called Flash surveys) were conducted in France to estimate the variant circulation in the country and at regional level at different moments in time. Flash 1 survey^4,5^ was conducted on January 7-8. It analyzed through Thermo Fisher 11,916 PCR positive samples out of 183,363 samples collected on January 7-8 by participating laboratories. 298 samples were confirmed B.1.1.7 variant, corresponding to 70% of analyzed S-gene dropouts, leading to 3.3% of new cases on January 7-8 due to SARS-CoV-2 B.1.1.7 variant in France. The estimated proportion of B.1.1.7 in Île-de-France and Nouvelle Aquitaine was 6.9% and 1.7%, respectively^5^.

Flash2 survey^6^ was conducted on January 27, and preliminary estimates are available at this time, as sequencing is still ongoing. 10,152 PCR samples were found positive out of 119,333 samples collected on January 27 by participating laboratories. S-gene target failure was identified in 474 cases out of 3,601 positive cases analyzed through Thermo Fisher, leading to an estimated 13.2% of S-gene target failure in France, 20.3% in Île-de-France and 12.6% in Nouvelle-Aquitaine. As genome sequencing is still undergoing, we use these data and estimate B.1.1.7 penetration as a 70% to 100% proportion of S-gene target failure percentages of positive tests (positive predicted value, PPV), given that 70% was the proportion estimated in the Flash1 survey. We considered that B.1.1.7 frequency observed in the surveys is described by a beta-binomial distribution to allow for dispersion due to spatial heterogeneity, and provide a 95% confidence interval for each PPV. Assuming 100% PPV, estimated median and 95% confidence intervals of B.1.1.7 penetration are: 13.2% [11.7, 14.8]% in France; 20.3 [17.1, 23.7]% in Île-de-France; 12.6% [7.3, 19.1]% in Nouvelle Aquitaine. Assuming 70% PPV would instead lead to: 9.2% [8.0, 10.6]% in France; 14.2% [11.5, 17.3]% in Île-de-France; 8.8% [4.5, 14.6]% in Nouvelle Aquitaine. These estimates are plotted in Figure 2 of the manuscript.

Starting w06, genomic surveillance changed into screening for specific mutations to identify suspect B.1.1.7 (and other variants^7^). The proportion of B.1.1.7 variant among screened samples that resulted positive to PCR are reported in **Table S1**.

For each estimate, we provide in **Table S1** here below and **Figure 2** of the main text a confidence interval assuming B.1.1.7 frequency is described by a beta-binomial distribution to allow for dispersion due to spatial heterogeneity and variations in detection protocols.

**Table S1.**
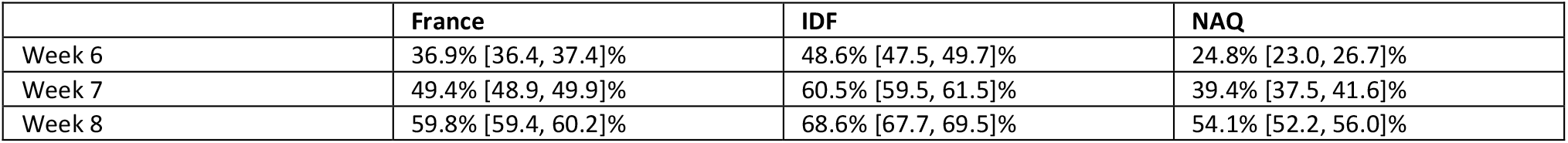
Proportion of suspect B.1.1.7.

## 2. SARS-CoV-2 two-strain transmission model

### 2.1. Compartmental model and parameters

We use a stochastic discrete age-stratified transmission model, integrating demographic, age profile, social contact data, mobility data, data on adoption of preventive measures, to account for age-specific behaviors over time and role in COVID-19 transmission. Four age classes are considered: [0-11), [11-19), [19-65), and 65+ years old (children, adolescents, adults, seniors). Transmission dynamics follows a compartmental scheme specific for COVID-19 (**Figure S1**) where individuals are divided into susceptible, exposed, infectious, hospitalized and recovered. The infectious phase is divided into two steps: a prodromic phase (Ip) and a phase where individuals may remain either asymptomatic (Ias, with probability p_a_=40%^8^) or develop symptoms. In the latter case, we distinguished between different degrees of severity of symptoms (paucisymptomatic (I_ps_), individuals with mild symptoms (I_ms_), or severe symptoms (I_ss_) requiring hospitalization^3,9,10^). Prodromic, asymptomatic and paucisymptomatic individuals have a reduced transmissibility^11^. A reduced susceptibility was considered for children and adolescents, along with a reduced relative transmissibility of children based on available evidence^12–17^. We assume that infectious individuals with severe symptoms reduce of 75% their number of contacts because of the illness they experience^18^. Parameter values are reported in **Table S2**.

Sensitivity analysis on the probability of becoming symptomatic and the transmissibility of children was performed in previous work^1,2,19^.

**Figure S1.**
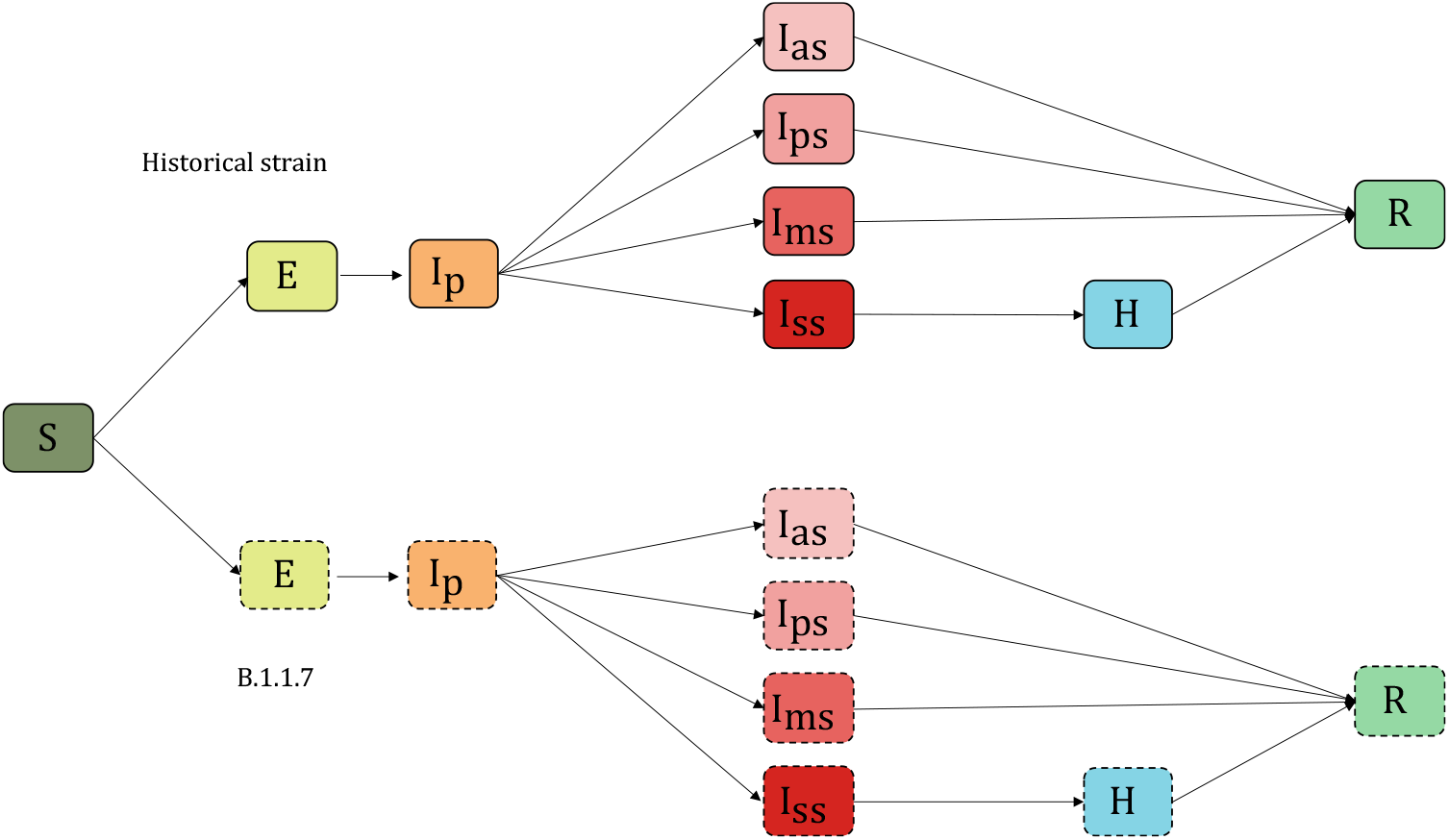
Two-strains compartmental scheme. Compartments with continuous line (top) account for the diffusion of historical strain, compartments with dashed line (bottom) account for the diffusion of B.1.1.7 variant. S=Susceptible, E=Exposed, I_p_= Infectious in the prodromic phase, I_as_=Asymptomatic Infectious, I_ps_=Paucysymptomatic Infectious, I_ms_=Symptomatic Infectious with mild symptoms, I_ss_=Symptomatic Infectious with severe symptoms, H=severe case admitted to the hospital, R=recovered.

**Table S2.** Parameters, values, and sources used to define the compartmental model.

| Variable | Description | Value | Source |
| --- | --- | --- | --- |
| $\theta^{-1}$ | Incubation period | 5.2d | 20 |
| $\mu_p^{-1}$ | Duration of prodromal phase | 1.5d, computed as the fraction of pre-symptomatic transmission events out of pre-symptomatic plus symptomatic transmission events. | 21 |
| $\epsilon^{-1}$ | Latency period | $\theta^{-1} - \mu_p^{-1}$ | - |
| $p_a$ | Probability of being asymptomatic | 0.4 | 8 |
| $p_{ps}$ | If symptomatic, probability of being paucisymptomatic | 1 for children, adolescents<br>0.2 for adults, seniors | 9 |
| $p_{ms}$ | If symptomatic, probability of developing mild symptoms | 0 for children, adolescents<br>0.7704 for adults<br>0.546 for seniors | 3,9,10 |
| $p_{ss}$ | If symptomatic, probability of developing severe symptoms | 0 for children, adolescents<br>0.0296 for adults<br>0.254 for seniors | 3,10 |
| $g$ | Generation time | 6.6d | 22 |
| $\mu^{-1}$ | Infectious period | 2.3d, chosen accordingly to generation time distribution | - |
| $r_\beta$ | Relative infectiousness of $I_p, I_a, I_{ps}$ | 0.25 for children<br>0.55 for adolescents, adults, seniors | 11 |
| $s$ | Relative susceptibility | 0.5 for children, adolescents<br>1 for adults, seniors | 13 |

### 2.2. Generation time distribution

The generation time distribution was computed based on the approach of Ref.^23^. Let *X* and *Y* be the random variables describing the latency period and the infectious period, respectively. Then the distribution of the generation time is the result of the convolution *g* * *h*_*S*_, with *g* being the probability density function of *X* and

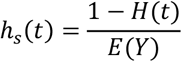

where *H* is the cumulative distribution function of *Y*, and *E*(*Y*)is the mean.

In the compartmental model under consideration (**Figure S1**), we have that *X* is exponentially distributed with rate *ϵ*, and *Y* is the sum of two exponentially distributed random variables (prodromic phase and infectious period, with rate *μ*_*p*_ and *μ* respectively). Computations show that the corresponding generation time distribution is

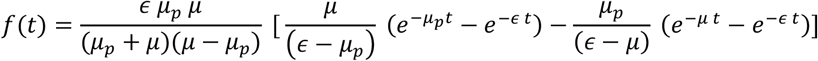

Given the values of *ϵ* and *μ*_*p*_ informed from the literature (**Table S2**), we chose *μ* so that the mean of the generation time equals to 6.6 days. The shape of the distribution is displayed in **Figure S2** and it closely resembles a gamma distribution with mean 6.6 and shape parameter 1.87, estimated in Ref^22^.

**Figure S2.**
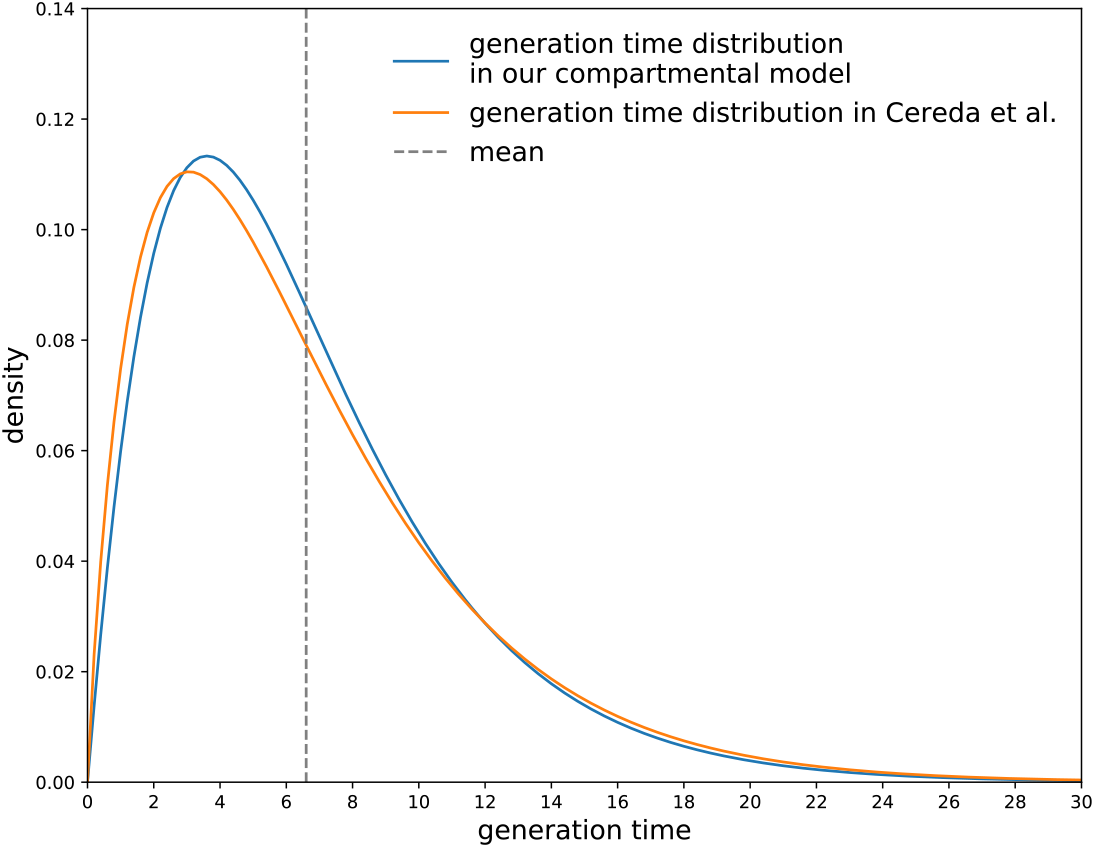
Distribution of the generation time. The generation time distribution corresponding to our compartmental model (blue) in comparison with the distribution estimated in Ref.^22^ (orange).

### 2.3. Parameterization of contact matrices from empirical data

Social mixing was informed from behavioral data and was modeled through the parametrization of contact matrices. In particular we considered attendance at school^24^, percentage of telework^25^, and adoption of physical distancing over time^26^.

Contacts at school were considered according to the school calendar. In France all schools are in session with 100% physical presence since the start of the school calendar in September. In the period of May-July, after the first lockdown, schools were open but attendance was on a voluntary basis^19^.

Social contacts at work were modified to account for the percentage of workers not going to their place of work over time, following the variation of presence at workplaces based on Google Mobility Trends^25^ (**Figure S3**).

To account for individuals’ risk protection behavior over time, we parametrized contact matrices with the percentage of population avoiding physical contacts from the results of regular large-scale surveys conducted by Santé Publique France (CoviPrev^26^). From these data, we also estimated that seniors have a higher risk aversion behavior compared to other age classes, leading to an average additional 30% reduction of their physical contacts^1^.

The contacts in leisure and non-essential activities were informed based on implemented restrictions and mobility data in community settings (see e.g. use of transport and visits to retail in **Figure S4**). A sensitivity analyses on contacts in leisure and non-essential activities was conducted in Ref.^1^.

**Figure S3.**
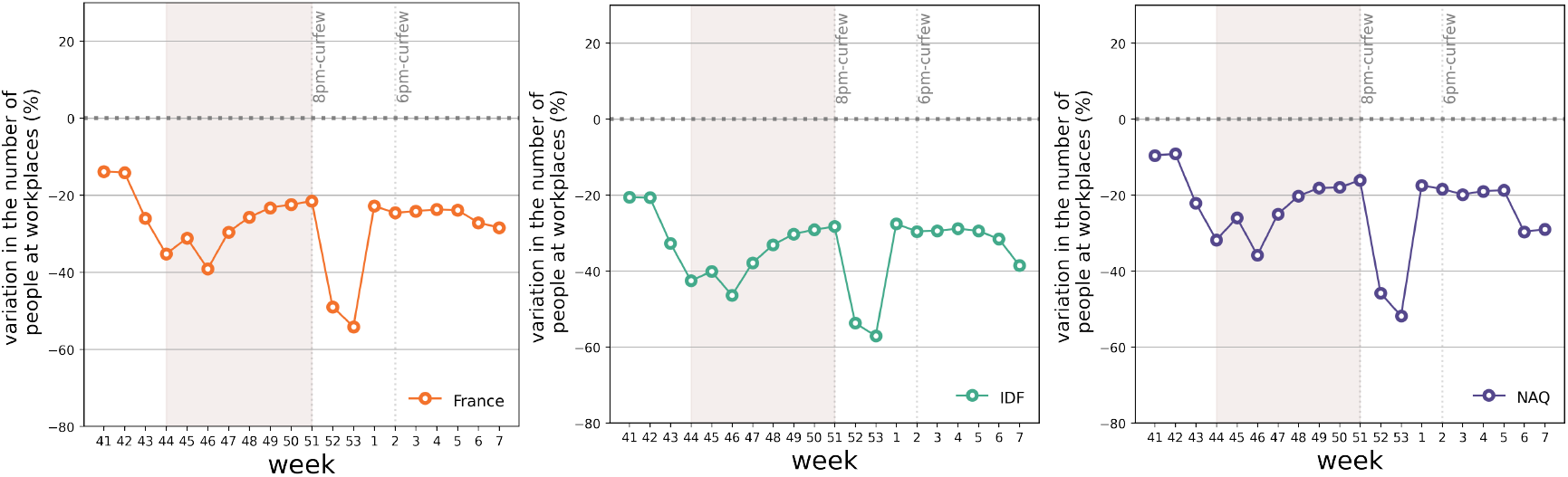
Estimated change in presence at workplace locations over time and by region based on Google location history data^25^.

**Figure S4.**
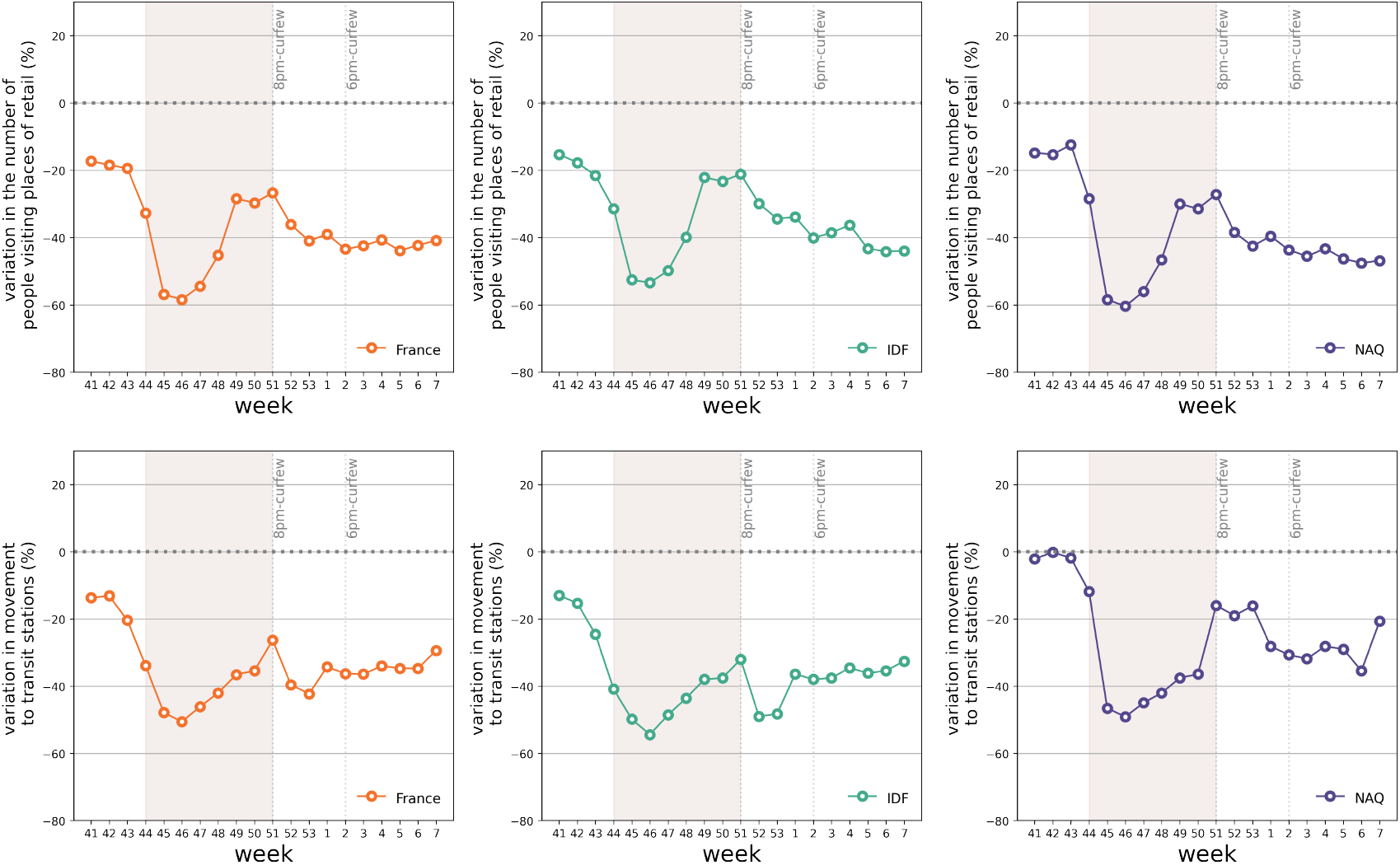
Estimated change in the number of people visiting places of retail (top) and in the movement to transit stations (bottom) over time and by region based on Google location history data^25^.

In prior work^1^, we compared our approach where the contact matrix is parameterized with the various data sources described above with a simplified version of the model that neglects these input data. This version assumes that all changes in the epidemic trajectory are absorbed exclusively by the transmissibility per contact. This is equivalent to normalize the contact matrix to its largest eigenvalue and estimate the reproductive ratio over time, as also done in other works^3^. Results showed that our model better describes the observed trajectories, thus indicating that changes in age-stratified contact patterns are important to capture the epidemic dynamics.

The code for the model is available at the link provided in Ref.^1^.

### 2.4. B.1.1.7 variant

We considered the co-circulation of B.1.1.7 variant together with the historical strain. Complete cross-immunity and 60% increased transmissibility (range 50%-70%)^5,27,28^ were assumed for the B.1.1.7 variant compared to the historical strain (**Figure S1)**. In the results considered in the main paper, we did not consider further differences between the two strains (generation time, hospitalization or severity rate). Recent results confirm that infection with lineage B.1.1.7 was associated with an increased risk of hospitalization compared with other lineages (adjusted OR of 1.64 (95%CI, 1.32-2.04))^29^. For sensitivity, we considered a 64% increase in hospitalization rates if infected with B.1.1.7 and refitted the model to hospitalization data to provide updated projections under this condition. Results are shown in section 4.

B.1.1.7 variant was initialized on January 7, 2021 (in w01) using the estimates of the first large-scale nationwide genomic surveillance survey (Flash1, see section 1 and main text). No other information was assumed, beyond the increased transmissibility advantage (and the increased hospitalization rate, for sensitivity). As such, no specific dynamics on the two strains is imposed *a priori*, and the trajectories predicted by the model are the result of the fit to hospitalization data (see next section).

### 2.5. Vaccination rollout campaign

Following estimated plans for rolling out the vaccination campaign, we simulated a rollout scenario based on the administration of 100,000 doses per day in France from w04, prioritized to the older age class. This was based on recent data, reporting an average daily rhythm of vaccination of 99,500 doses (including first and second doses) from w04 to w08^30^. We considered 75% vaccine efficacy against susceptibility^31^, 65% vaccine efficacy against transmission^32^, and a range between 40% and 80% for vaccine efficacy against symptoms given infection, computed from the estimated vaccine reduction of symptomatic disease^32,33^, between 85%^31^ and 95%^34,35^. Estimates were found to be similar when evaluated 14 days after the first dose or 1-2 weeks after the second dose^35^, therefore we assumed efficacy to start 14 days after the first injection. We also considered for sensitivity: (i) a reduced efficacy against transmission (50%); (ii) no efficacy against susceptibility and 90% efficacy against symptoms.

Following the announcements by French authorities on March 4 aiming at administering 10 million first doses till mid-April^36^, we considered an acceleration in the daily rhythm in w10 (starting March 8) with 200,000 doses per day (only first doses). For sensitivity, we considered a more optimistic rollout of 300,000 doses/day (only first doses; this latter estimate based on observations that 250,000 doses were administered on March 5 after the announcements^37^). These rollouts are compared to a stable rhythm of 100,000 doses (used for first and second injections).

## 3. Inference framework

Once the model is parameterized with the data described above, we infer the transmission rate per contact by fitting the model to daily hospital admission data through a maximum likelihood procedure in each pandemic phase. More precisely, prior to lockdown and in absence of intervention (period January-March 2020), we estimated {*β, t*_0_} where *β* is the transmission rate per contact and *t*_0_ the date of the start of the simulation, seeded with 10 infectious individuals. Then, in each phase we estimated *α*_*phase*_, i.e. the scaling factor of the transmission rate per contact specific to the pandemic phase under study (e.g. lockdown, exit from lockdown, summer, start of second wave, second lockdown, etc.). The transmission rate per contact in each phase is then defined as the transmission rate per contact in the pre-lockdown phase *β* multiplied by the scaling factor *α*_*phase*_. A pandemic phase is defined by the interventions implemented (e.g. lockdown, curfew, and other restrictions) and activity of the population (school holidays, summer holidays, etc.).

We used simulations of the stochastic model to predict values for all quantities of interest (500 stochastic simulations each time). We fitted the model to the daily count of hospitalizations *H*_*obs*_(*d*)on day *d*. The likelihood function is of the form

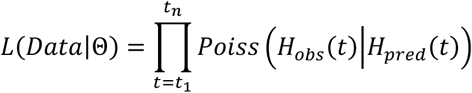

where Θ indicates the set of parameters to be estimated, *H*_*obs*_(*t*)is the observed number of hospital admissions on day *t, H*_*pred*_(*t*)is the number of hospital admissions predicted by the model on day *t, poiss* (⋅|*H*_*pred*_(*t*)) is the probability mass function of a Poisson distribution with mean *H*_*pred*_(*t*), and [*t*_1_, *t*_*n*_] is the time window considered for the fit. The effect of social distancing in January 2021 was estimated in the w02-w05 period, to account for the expected delay from the implementation of the measures (starting at the end of week 53, then progressively strengthened till w02) and hospitalizations.

Wald confidence intervals for the scaling factor *α* were computed by fitting a quadratic function on the loglikelihood values around the MLE, to estimate Fisher’s information.

In prior work^1^ we showed that the stochasticity of the model is the main source of uncertainty in the predictions.

Simulations are initialized with 10 infected adults in the *I*_p_ compartment at the estimated time *t*_0_ and progress over the entire 2020 to describe the full trajectory of the epidemic in France (and in each region under study) and build up the immunity in the population, prior to the arrival of B.1.1.7 variant. The model was validated against the estimates of three independent serological surveys conducted in France^1^. The second wave, under the effect of the second national lockdown, is shown in the Figures reporting the trajectory (main text and SI) to provide the epidemic context of COVID-19 pandemic in France, and a reference frame for the critical levels of hospitalizations that triggered the second lockdown.

## 4. Additional results and sensitivity analysis

### 4.1. Impact of B.1.1.7 transmissibility advantage

**Figure S5.**
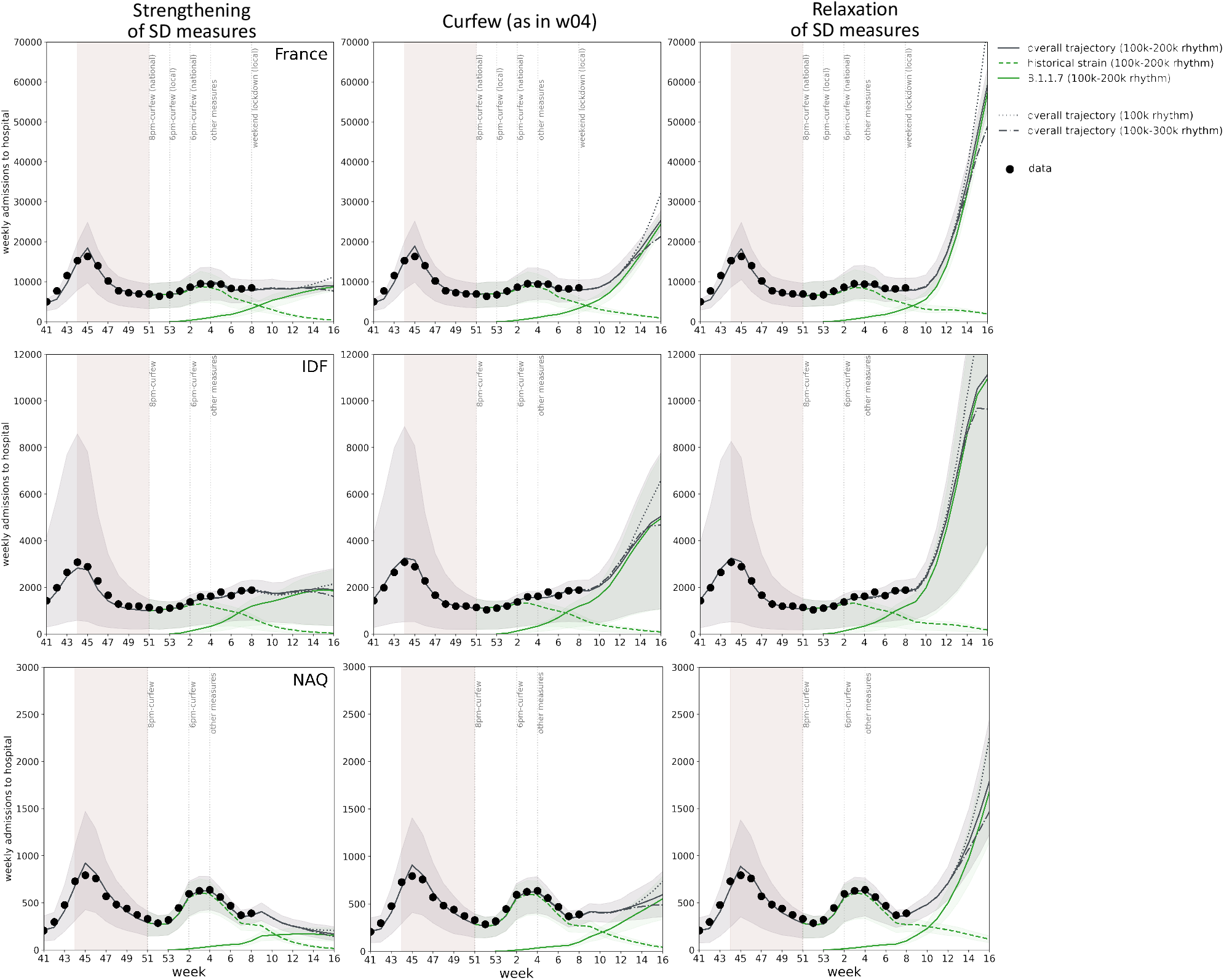
Impact of 50% transmissibility increase on the projected weekly hospitalizations due to SARS-CoV-2 historical strain and B.1.1.7 variant. From left to right, different scenarios considered after winter school holidays: strengthening of social distancing (SD) measures scenario, equivalent to the second lockdown; curfew scenario, estimated in w04 and assuming no additional changes; relaxation of SD measures scenario, compatible with the situation at the start of the year before increased restrictions. From top to bottom: France, Île-de-France (IDF), Nouvelle Aquitaine (NAQ). The solid grey curve refers to the median overall trajectory, obtained under the accelerated vaccination rollout (100k-200k doses/day) and due to the concurrent circulation of the historical strain (dashed green curve) and B.1.1.7 variant (solid green curve), assuming 50% increase in transmissibility. A slower (100k, dotted curve) and optimistic (100k-300k, dot-dashed curve) vaccination rhythms are also shown (only median curves of the overall trajectories are shown, for the sake of visualization). The shaded area around the curves corresponds to the 95% probability range obtained from 500 stochastic simulations. Dots correspond to weekly hospital admission data. The model is fit to daily hospital admissions since the start of the epidemic, propagating uncertainty over time; the figure shows weekly data to simplify the visualization. The second wave is shown for reference, together with indications of the timing of social distancing measures; the shaded rectangle around the second wave corresponds to the second lockdown.

**Figure S6.**
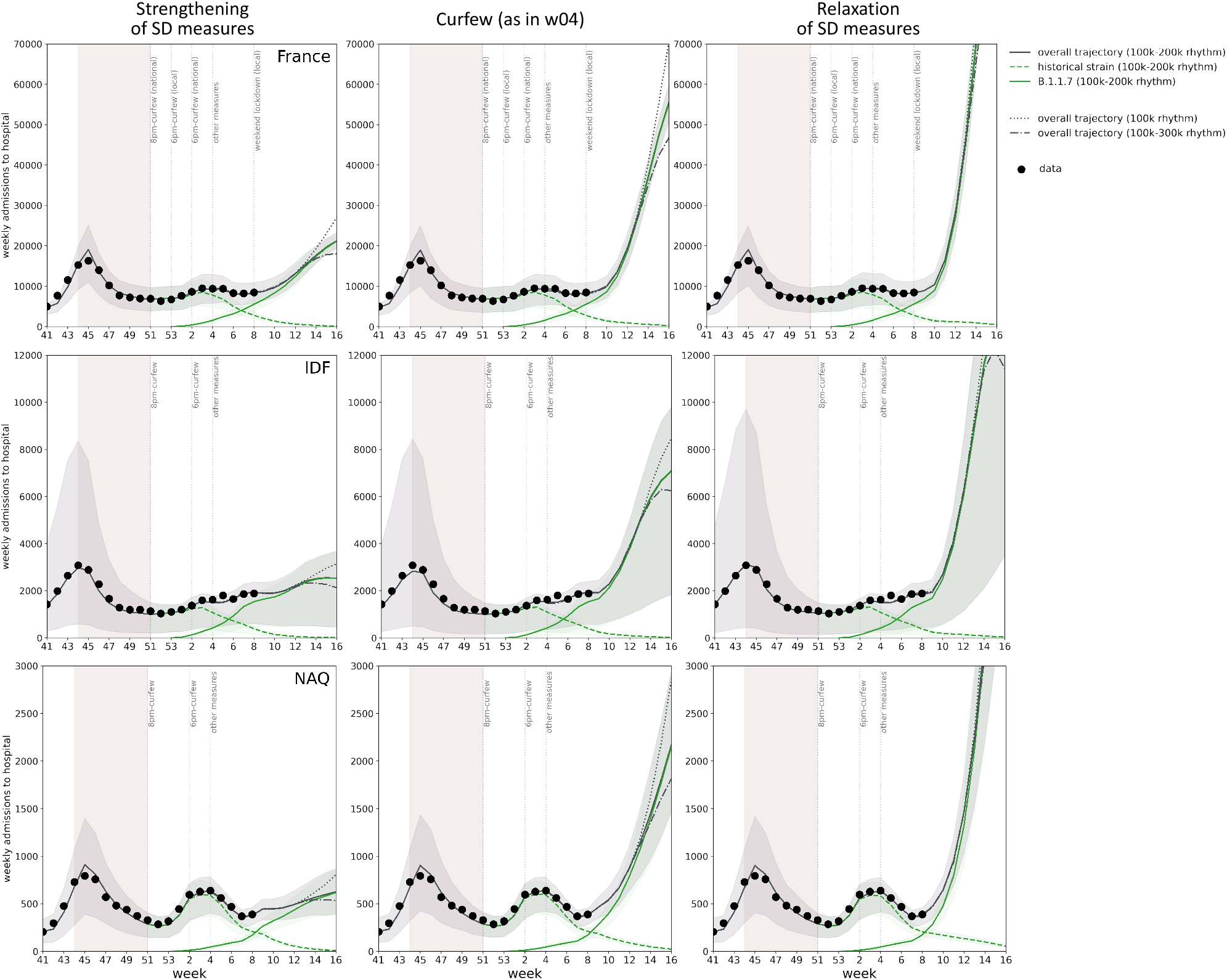
Impact of 70% transmissibility increase on the projected weekly hospitalizations due to SARS-CoV-2 historical strain and B.1.1.7 variant. From left to right, different scenarios considered after winter school holidays: strengthening of social distancing (SD) measures scenario, equivalent to the second lockdown; curfew scenario, estimated in w04 and assuming no additional changes; relaxation of SD measures scenario, compatible with the situation at the start of the year before increased restrictions. From top to bottom: France, Île-de-France (IDF), Nouvelle Aquitaine (NAQ). The solid grey curve refers to the median overall trajectory, obtained under the accelerated vaccination rollout (100k-200k doses/day) and due to the concurrent circulation of the historical strain (dashed green curve) and B.1.1.7 variant (solid green curve), assuming 70% increase in transmissibility. A slower (100k, dotted curve) and optimistic (100k-300k, dot-dashed curve) vaccination rhythms are also shown (only median curves of the overall trajectories are shown, for the sake of visualization). The shaded area around the curves corresponds to the 95% probability range obtained from 500 stochastic simulations. Dots correspond to weekly hospital admission data. The model is fit to daily hospital admissions since the start of the epidemic, propagating uncertainty over time; the figure shows weekly data to simplify the visualization. The second wave is shown for reference, together with indications of the timing of social distancing measures; the shaded rectangle around the second wave corresponds to the second lockdown.

**Figure S7.**
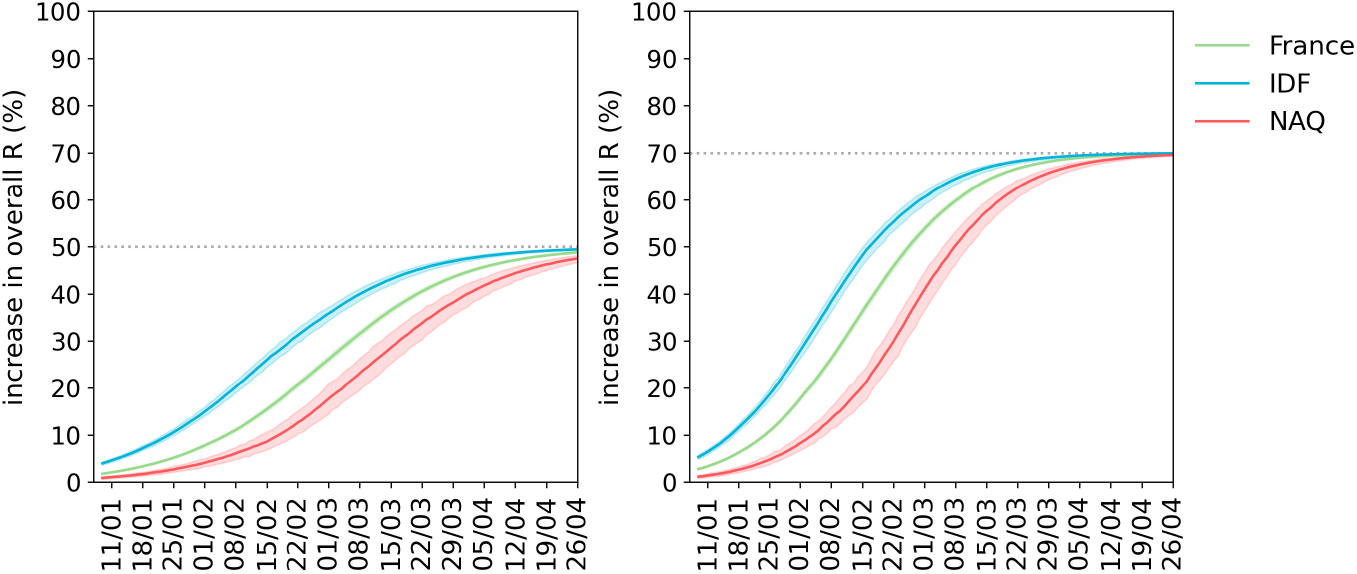
Percentage increase in the overall effective reproductive number at the population level due to the increased penetration of the variant. Results are shown for 50% transmissibility increase (left panel) and 70% transmissibility increase of the B.1.1.7 strain (right panel). Curves represent median values for France (green), Île-de-France (blue), Nouvelle Aquitaine (red); the shaded area around the curves corresponds to the 95% probability range obtained from 500 stochastic simulations.

### 4.2. Impact of strengthening / relaxation of social distancing measures on projected B.1.1.7 prevalence

**Figure S8.**
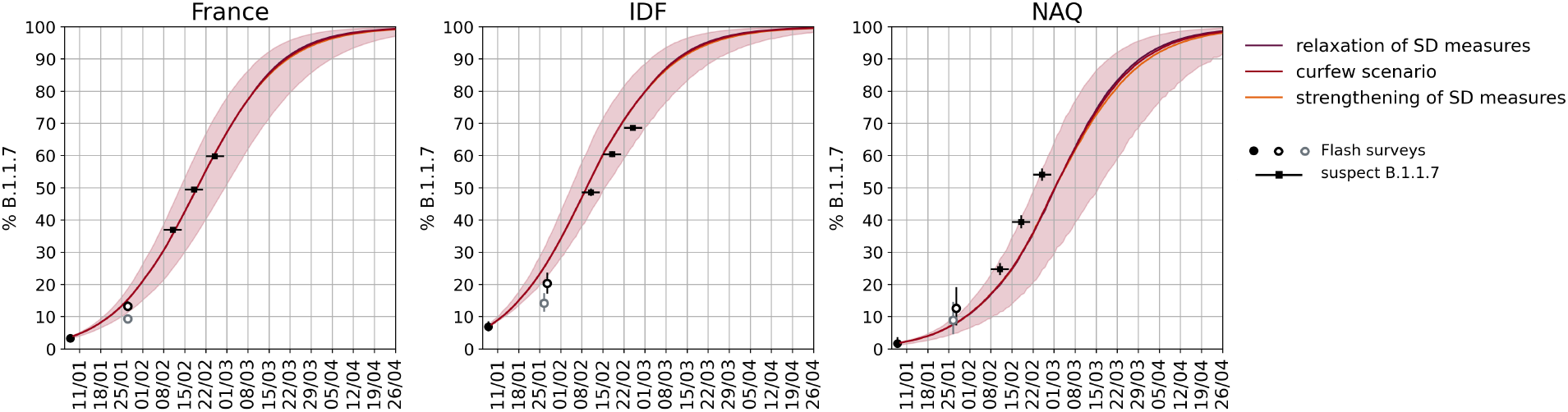
Impact of strengthening and relaxation of SD measures on the B.1.1.7 projected prevalence over time. Estimated percentage of B.1.1.7 cases over time, assuming 60% (50-70%) increase in transmissibility for the variant. From left to right: mainland France, Île-de-France region (IDF) and Nouvelle Aquitaine region (NAQ). Circles represent the estimates from Flash surveys. As sequencing is still ongoing, estimates from the second Flash survey are based on S-gene target failure percentages of positive tests for each territory and positive predicted value between 70% (grey border circle, national estimate from Flash1) and 100% (black border circles). Weekly data on suspect B.1.1.7 are reported with black squares; the horizontal line correspond to the week of reference. 95% confidence interval are estimated assuming a beta-binomial distribution to allow for dispersion due to spatial heterogeneity and variations in detection protocols. Curves represent median values; the shaded area around the curves corresponds to the 95% probability range obtained from 500 stochastic simulations. Color shade refers to the curfew scenario (intermediate color) and the scenarios with strengthening (lighter color) and relaxation (darker color) of social distancing measures. Results show that social distancing measures of this intensity do not have a significant impact on the prevalence of B.1.1.7 over time, as they concurrently act on both strains.

### 4.3. Impact of more restrictive social distancing measures on projected trends of hospital admissions

Here we present the results obtained under more restrictive measures, corresponding to a 20% reduction of the effective reproductive number estimated for the curfew in w04. Such reduction is estimated to be necessary to avoid a considerable increase in hospital admissions in France, and achieve a reduction in Île-de-France.

**Figure S9.**
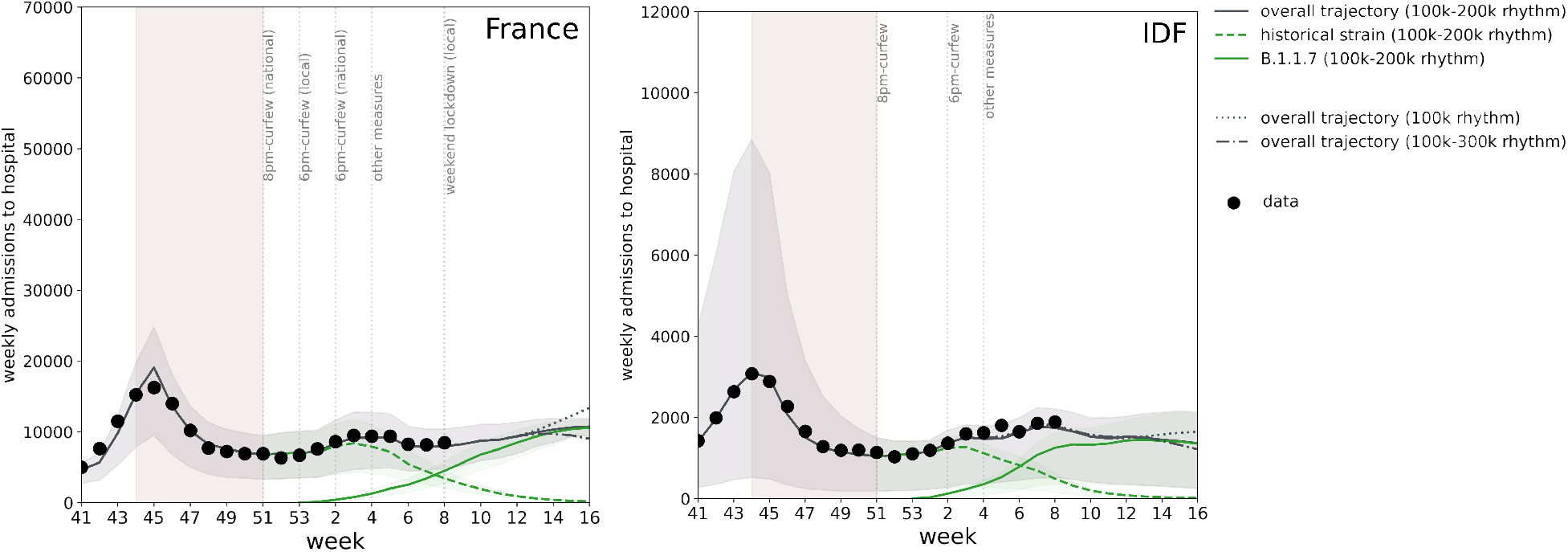
Impact of 20% reduction of the effective reproductive number on the projected weekly hospitalizations due to SARS-CoV-2 historical strain and B.1.1.7 variant. From left to right: mainland France, Île-de-France (IDF). Scenario considered after winter school holidays: strengthening of social distancing (SD) measures corresponding to a reduction of 20% of the effective reproduction number. The solid grey curve refers to the median overall trajectory, obtained under the accelerated vaccination rollout (100k-200k doses/day) and due to the concurrent circulation of the historical strain (dashed green curve) and B.1.1.7 variant (solid green curve), assuming 60% increase in transmissibility. A slower (100k, dotted curve) and optimistic (100k-300k, dot-dashed curve) vaccination rhythms are also shown (only median curves of the overall trajectories are shown, for the sake of visualization). The shaded area around the curves corresponds to the 95% probability range obtained from 500 stochastic simulations. Dots correspond to weekly hospital admission data. The model is fit to daily hospital admissions since the start of the epidemic, propagating uncertainty over time; the figure shows weekly data to simplify the visualization. The second wave is shown for reference, together with indications of the timing of social distancing measures; the shaded rectangle around the second wave corresponds to the second lockdown.

### 4.4. Sensitivity analysis on vaccine daily rhythm and efficacy

#### 4.4.1. Impact of vaccination rhythms on the projected weekly hospitalizations

**Figure S10** shows the effect of vaccination under different rhythms compared to a no vaccination scenario for France, and **Table S3** shows the expected week at which hospitalizations would reach the levels of the second wave, under the different social distancing scenarios and vaccination rhythms, for all territories.

**Figure S10.**
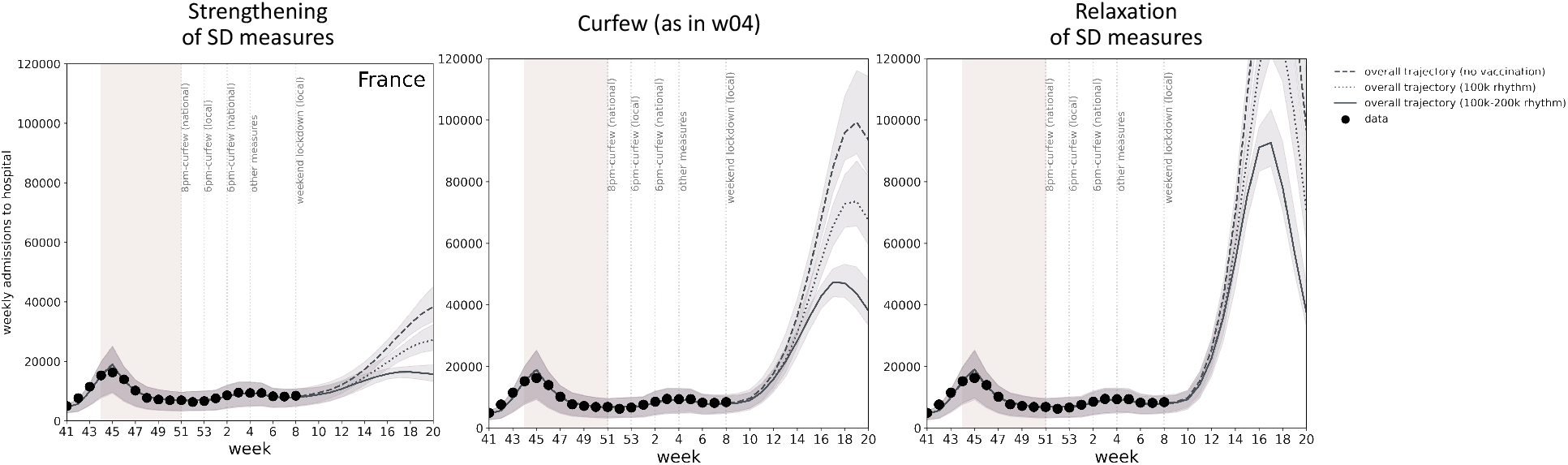
Impact of vaccination rhythms on the projected weekly hospitalizations due to SARS-CoV-2 historical strain and B.1.1.7 variant. Results are shown for mainland France. From left to right, different scenarios considered after winter school holidays: strengthening of social distancing (SD) measures scenario, equivalent to the second lockdown; curfew scenario, estimated in w04 and assuming no additional changes; relaxation of SD measures scenario, compatible with the situation at the start of the year before increased restrictions. Dots correspond to weekly hospital admission data. Curves refer to the expected trajectory with the accelerated vaccination rhythm (100k-200k doses/day, solid curve, as in the main paper), stable rhythm (100k doses/day, dotted), and no vaccination (dashed curve). The shaded area around the curves corresponds to the 95% probability range obtained from 500 stochastic simulations. Dots correspond to weekly hospital admission data. The model is fit to daily hospital admissions since the start of the epidemic, propagating uncertainty over time; the figure shows weekly data to simplify the visualization. The second wave is shown for reference, together with indications of the timing of social distancing measures; the shaded rectangle around the second wave corresponds to the second lockdown

**Table S3.** Estimated week at which hospitalizations exceed the peak value of the second wave in France and in the two regions under study, for varying vaccination rollouts. Projections after school holidays consider the curfew scenario (estimated before school holidays, assuming no additional interventions, central column), a scenario assuming a strengthening of social distancing (SD) measures (left column), and one assuming a relaxation of social distancing measures (right column). Results correspond to 60% increase of transmissibility of the variant. Results cannot integrate yet the effect of the weekend lockdowns in certain areas, nor Easter school holidays. Ranges refer to 95% probability ranges; “--” indicates that the peak level is not reached. Figures of daily administration of vaccine doses in the table refer to the national level. In each region daily rhythms are computed considering a population-weighted distribution for the senior age class.

|  |  | Peak weekly hospitalizations of second wave |  |  |  |
| --- | --- | --- | --- | --- | --- |
|  |  | Vaccine doses per day from w10 | Strengthening of SD measures | Curfew (as in w04) | Relaxation of SD measures |
| France | 17,000 weekly hospitalizations | 100k rhythm | week 15 (14-16) | week 13 (12-13) | week 12 (11-12) |
|  |  | 200k rhythm | week 16 (16-17) | week 13 (12-14) | week 12 (11-13) |
|  |  | 300k rhythm | week 16 (16-17) | week 13 (12-13) | week 12 (11-12) |
| Île-de-France | 3,000 weekly hospitalizations | 100k rhythm | -- | week 12 (11-14) | week 11 (10-14) |
|  |  | 200k rhythm | -- | week 12 (11-14) | week 11 (10-13) |
|  |  | 300k rhythm | -- | week 12 (11-12) | week 11 (10-12) |
| Nouvelle Aquitaine | 800 weekly hospitalizations | 100k rhythm | After week 16 | week 14 (12-15) | week 12 (11-13) |
|  |  | 200k rhythm | -- | week 14 (12-16) | week 12 (11-13) |
|  |  | 300k rhythm | -- | week 14 (13-18) | week 12 (11-13) |

#### 4.4.2. Impact of vaccine efficacy on the projected weekly hospitalizations

**Figure S11** reports the sensitivity on vaccine efficacy.

**Figure S11.**
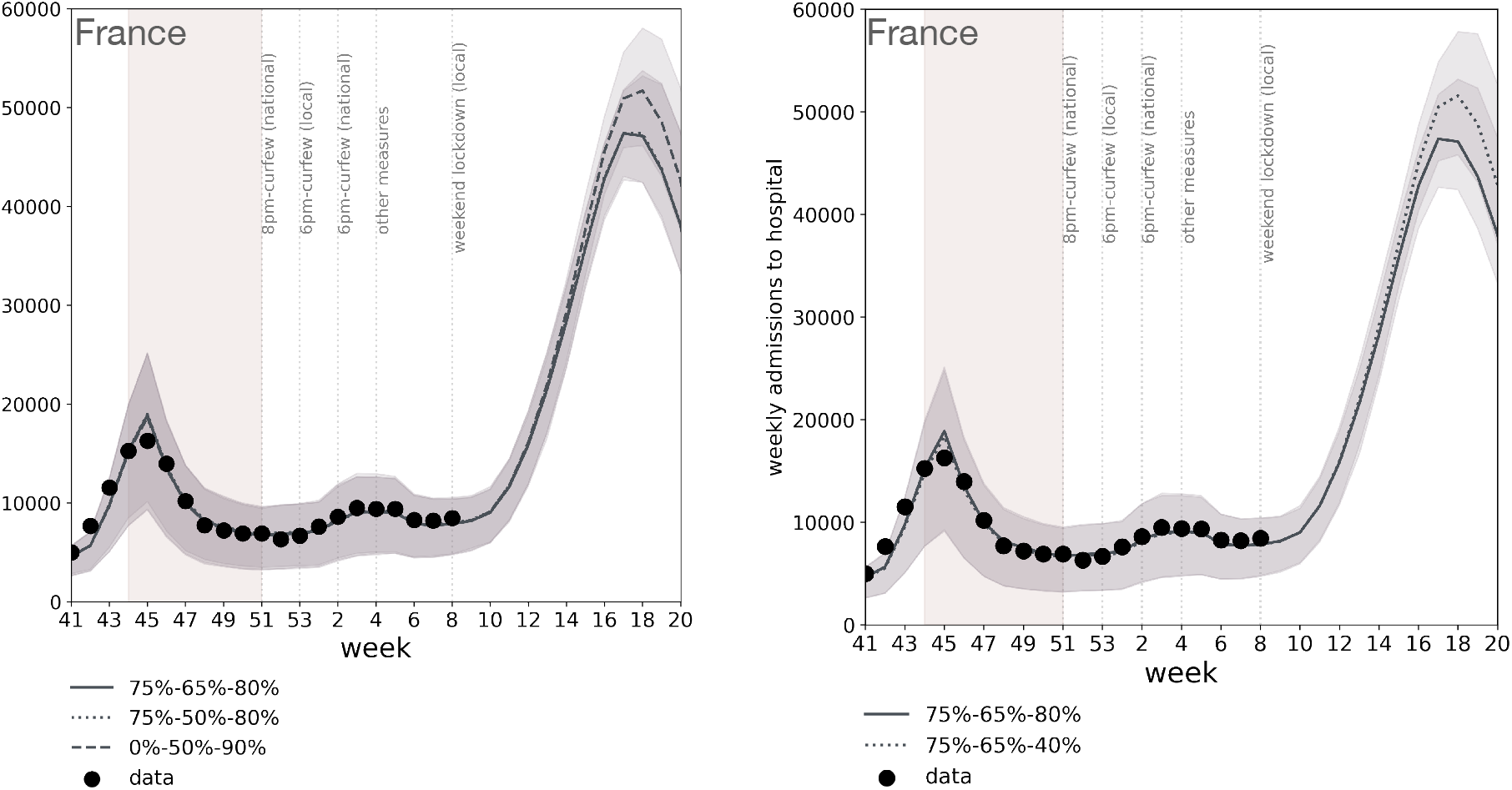
Impact of vaccine efficacy on the projected weekly hospitalizations due to SARS-CoV-2 historical strain and B.1.1.7 variant. Results are shown for mainland France. Scenario considered after winter school holidays: curfew scenario, estimated in w04 and assuming no additional changes. Curves refer to the median overall trajectory, obtained under the accelerated vaccination rollout (100k-200k doses/day) and due to the concurrent circulation of the historical strain and B.1.1.7 variant, assuming 60% increase in transmissibility. In both panels, solid line is obtained assuming a 75% vaccine efficacy against susceptibility, 65% vaccine efficacy against transmission and 80% vaccine efficacy against symptoms given infection, as in the main paper (indicated as 75%-65%-80% in the Figure legend). Left panel: dotted line is obtained assuming 75% vaccine efficacy against susceptibility, 50% vaccine efficacy against transmission and 80% vaccine efficacy against symptoms given infection (75%-50%-80%); dashed line is obtained assuming no efficacy against susceptibility, 50% vaccine efficacy against transmission, and 90% vaccine efficacy against symptoms given infection (0%-50%-90%). Right panel: dotted line is obtained assuming 75% vaccine efficacy against susceptibility, 65% vaccine efficacy against transmission and 40% vaccine efficacy against symptoms given infection (75%-65%-40%). Shaded area around the curves corresponds to the 95% probability range obtained from 500 stochastic simulations. Dots correspond to weekly hospital admission data. The model is fit to daily hospital admissions since the start of the epidemic, propagating uncertainty over time; the figure shows weekly data to simplify the visualization. The second wave is shown for reference, together with indications of the timing of social distancing measures; the shaded rectangle around the second wave corresponds to the second lockdown.

We note that our results may be an underestimation of the impact of vaccination on the epidemic trajectory as in our model priority is given to 65+ whereas vaccination is currently being rolled out first in the 75+ age class, characterized by a higher hospitalization rate. As our model does not break down further age classes above 65 years of age, vaccination cannot account for the higher advantage in targeting older age classes. On the other hand, we optimistically assume that vaccine efficacy is reached 2 weeks after the first injection, which was observed so far only for mRNA-1273 vaccine^35^.

### 4.5. Impact of increased hospitalization rates associated to B.1.1.7 infection

The increased hospitalization rate (+64%) after B.1.1.7 infection, recently estimated in Denmark^29^, is expected to lead to a higher peak of hospital admissions starting at the end of April (**Figure S12**), if curfew measures only are in place. Hospital admission would be however largely higher than the levels of the first and second peak, even with the considered vaccination rhythms.

**Figure S12.**
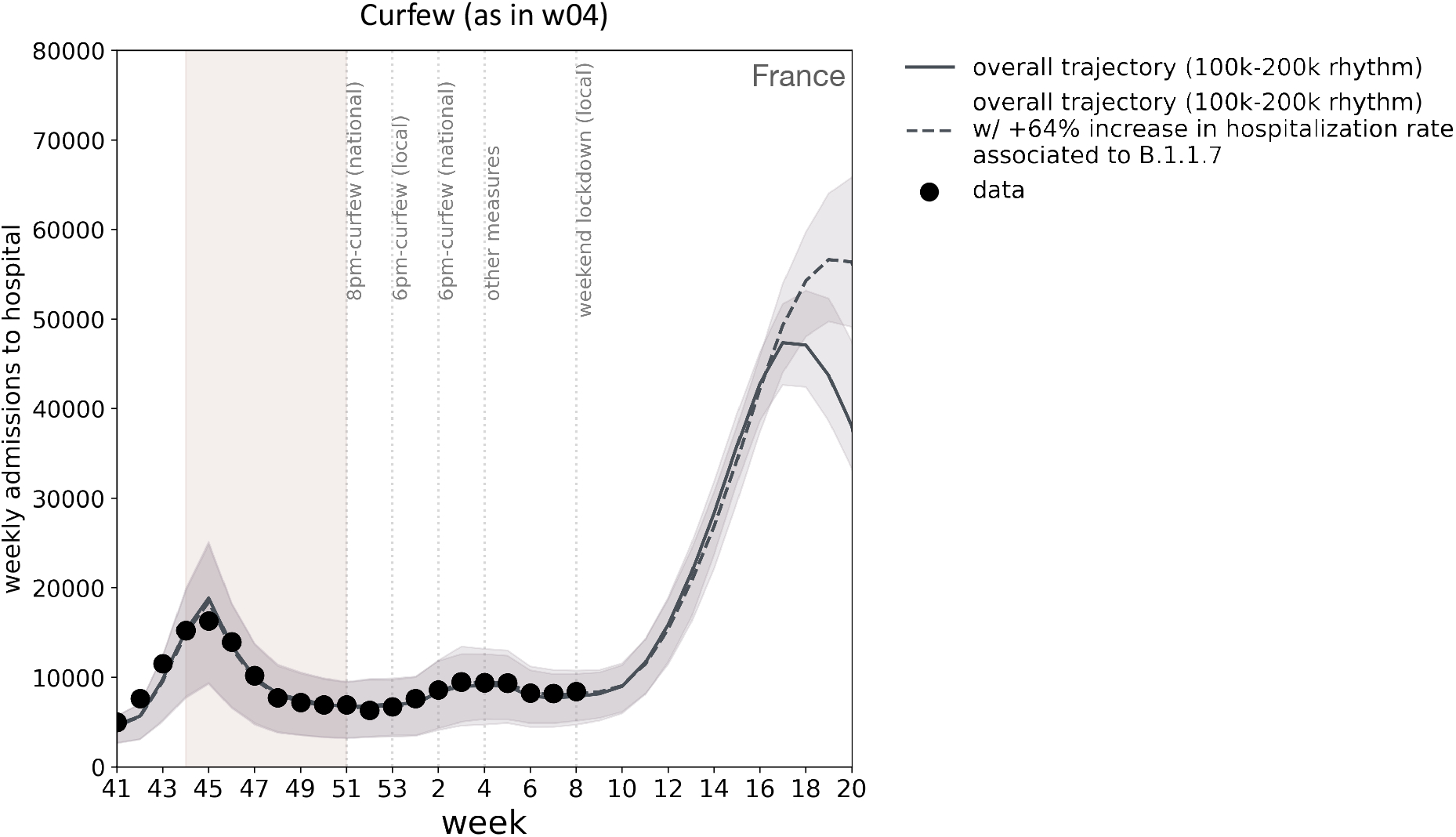
Impact of an increased hospitalization rate after B.1.1.7 infection on the projected weekly hospitalizations due to SARS-CoV-2 historical strain and B.1.1.7 variant. Results are shown for mainland France. Scenario considered after winter school holidays: curfew scenario, estimated in w04 and assuming no additional changes. Curves refer to the overall trajectory, due to the concurrent circulation of the historical strain and of the B.1.1.7 variant, assuming 60% increase in transmissibility and under the accelerated vaccination rollout (100k-200k doses/day). The solid grey curve refers to the median overall trajectory, obtained assuming the same hospitalization rates for infection due to the historical train and B.1.1.7. variant (as in the main paper). The dashed grey curve refers to the median overall trajectory, obtained assuming an increased hospitalization rate (+64%) associated to B.1.1.7 infection^29^.The shaded area around the curves corresponds to the 95% probability range obtained from 500 stochastic simulations. Dots correspond to weekly hospital admission data. The model is fit to daily hospital admissions since the start of the epidemic, propagating uncertainty over time; the figure shows weekly data to simplify the visualization. The second wave is shown for reference, together with indications of the timing of social distancing measures; the shaded rectangle around the second wave corresponds to the second lockdown.

